# Functional brain network dynamics of brooding in depression: insights from real-time fMRI neurofeedback

**DOI:** 10.1101/2024.05.05.24306889

**Authors:** Saampras Ganesan, Masaya Misaki, Andrew Zalesky, Aki Tsuchiyagaito

## Abstract

**Background:** Brooding is a critical symptom and prognostic factor of major depressive disorder (MDD), which involves passively dwelling on self-referential dysphoria and related abstractions. The neurobiology of brooding remains under characterized. We aimed to elucidate neural dynamics underlying brooding, and explore their responses to neurofeedback intervention in MDD.

**Methods:** We investigated functional MRI (fMRI) dynamic functional network connectivity (dFNC) in 36 MDD subjects and 26 healthy controls (HCs) during rest and brooding. Rest was measured before and after fMRI neurofeedback (MDD-active/sham: n=18/18, HC-active/sham: n=13/13). Baseline brooding severity was recorded using Ruminative Response Scale - Brooding subscale (RRS-B).

**Results:** Four recurrent dFNC states were identified. Measures of time spent were not significantly different between MDD and HC for any of these states during brooding or rest. RRS-B scores in MDD showed significant negative correlation with measures of time spent in dFNC state 3 during brooding (r=-0.5, p= 1.7E-3, FDR-significant). This state comprises strong connections spanning several brain systems involved in sensory, attentional and cognitive processing. Time spent in this anti-brooding dFNC state significantly increased following neurofeedback only in the MDD active group (z=-2.09, p=0.037).

**Limitations:** The sample size was small and imbalanced between groups. Brooding condition was not examined post-neurofeedback.

**Conclusion:** We identified a densely connected anti-brooding dFNC brain state in MDD. MDD subjects spent significantly longer time in this state after active neurofeedback intervention, highlighting neurofeedback’s potential for modulating dysfunctional brain dynamics to treat MDD.

## Introduction

Rumination refers to repeatedly dwelling on negative self-referential thought patterns, events and experiences (Ehring & Watkins, 2008; Nolen-Hoeksema et al., 2008; Treynor et al., 2003). This cognitive process has been increasingly recognized as maladaptive and implicated in the maintenance and exacerbation of major depressive disorder (MDD) and other mood disorders (Bessette et al., 2020; Ehring & Watkins, 2008; Watkins, 2009a b, 2009b a). It is also considered a crucial element within the research domain criteria (RDoC) framework (Tozzi et al., 2020). Among its components, brooding - the passive tendency to dwell on abstract causes and consequences of one’s problems, symptoms and dysphoric mood (Treynor et al., 2003) - stands out for its strong association with increased risk and sustenance of depression and mood disorders (Lackner & Fresco, 2016; Treynor et al., 2003; Watkins, 2009a).

Emerging functional magnetic resonance imaging (fMRI) literature on the neurobiology of rumination have broadly implicated aberrations within default-mode network (DMN), salience network (SN) and central executive network (CEN) (Berman et al., 2011, 2014; Hamilton et al., 2015; Jacob et al., 2020; Mısır et al., 2023; Zhou et al., 2020). These networks are associated with self-referential and autobiographical thinking (Raichle, 2015), awareness and arousal (Menon & Uddin, 2010), and adaptive cognitive control (Dosenbach et al., 2007) respectively. A meta-analysis of task-fMRI studies investigating rumination found convergent increases of activation in dorsal anterior cingulate cortex (ACC), precuneus, superior temporal gyrus (STG) and other areas, such that the significant findings maximally overlapped with DMN subsystems that are relevant to repetitive and passive mental dwelling on past events, future scenarios, and feelings (Zhou et al., 2020). Similarly, a recent systematic review implicated increased FC of subgenual ACC, posterior cingulate cortex (PCC), medial prefrontal cortex (mPFC), and amygdala, among other regions in DMN, SN and CEN (Mısır et al., 2023). However, despite the higher clinical significance of brooding compared to other rumination subtypes, the neurobiology of brooding remains unclear. Increased brooding has been associated with varying neurobiological changes across the fMRI literature, such as reduced FC between amygdala and temporal pole in MDD and healthy samples (Satyshur et al., 2018), increased FC between PCC and subgenual ACC during rest in MDD and healthy samples (Berman et al., 2011), reduced variability of DLPFC activity in MDD (Philippi et al., 2022), increased FC within SN (particularly involving dorsal ACC) in young girls (Ordaz et al., 2017), increased FC between insula and hippocampal areas in healthy individuals (X. Li et al., 2022), and FC changes in the triple-network (i.e., DMN, SN and CEN) in MDD (Pisner et al., 2019). These observations suggest that brooding is likely supported by excessive self-directed thought, impaired regulation of negative emotional stimuli and disrupted flexibility to disengage from repetitive negative thinking and dysphoria.

Brain function is largely dynamic and context-dependent (Rabinovich et al., 2012). Dynamic time-varying FC can illuminate complex time-varying neural interactions underlying fluctuating cognitive states that are typically missed by static time-averaged FC estimations (Hutchison et al., 2013). Studies investigating dynamic FC in rumination and MDD have observed links to disrupted FC dynamics of the DMN, CEN and other networks, suggesting impaired neural communications associated with cognitive control, flexibility and self-referential processing. High variability (and low stability) of FC dynamics in DMN regions such as mPFC, hippocampus and PCC (Chen (••) & Yan (•超•), 2021; Kaiser et al., 2016; Kim et al., 2023; Kucyi & Davis, 2014) was associated with increased rumination and mind-wandering across MDD and healthy samples, and dynamic FC of dorsal mPFC was found to strongly predict rumination in MDD (Kim et al., 2023). Similarly, lower stability and shorter dwelling in dynamic FC states with positive FC of DMN, sensorimotor areas and subcortical regions have been associated with MDD pathology (Long et al., 2020; Wu et al., 2019). In contrast, higher stability and longer dwelling in dynamic FC states with positive FC in DMN and CEN (Yao et al., 2019) and higher activity in SN, somatomotor and attention networks (Javaheripour et al., 2023) have also been observed during resting-state in MDD. Despite the emerging efforts to characterize dissociable dynamic FC states of rumination and MDD broadly, there is a paucity of literature examining dynamic FC associated with brooding.

The goal of this study is to bridge the gap in our understanding of neurobiological underpinnings of brooding by comparing dynamic FC properties between resting-state and experimentally induced brooding condition across MDD subjects and healthy controls (HCs). This approach may also inform the development of interventions that target the neural dysfunction underlying pathological brooding, like real-time fMRI neurofeedback (Pindi et al., 2022), where individuals learn to modulate a specific brain function and associated behavior with guidance from real-time feedback of personalized fMRI brain activity.

The primary aim of our study was to identify dynamic FC states most relevant to brooding severity in MDD subjects and HCs. Specifically, we aimed to estimate whole-brain, time-varying dynamic functional network connectivity (dFNC) states associated with brooding and resting-state fMRI using a well-validated dynamic FC analysis technique (Allen et al., 2014; Sendi et al., 2022), and subsequently examine the association between key temporal indices (like time spent) of the identified dFNC states and baseline brooding scores (measured using Rumination Response Brooding subscale (RRS-B)). An additional exploratory aim involved examining the impact of real-time fMRI neurofeedback on the dynamics of brooding-related dFNC states, thereby expanding on findings from our previous double-blind, randomized, and sham-controlled, clinical trial of real-time fMRI neurofeedback and its effects on static FC associated with brooding (Misaki et al., 2020; Tsuchiyagaito et al., 2021, 2023).

We hypothesized that: (1) During brooding, compared to HCs, MDD subjects would show significant decreases in time spent and increases in temporal variability in distinct dFNC states with strong connections within and between various DMN (e.g., PCC, mPFC), CEN (e.g., DLPFC), SN (e.g., insula, dorsal ACC), and subcortical (e.g., hippocampus, thalamus) regions, building upon prior observations of brooding-related static FC (Berman et al., 2011; X. Li et al., 2022; Ordaz et al., 2017; Philippi et al., 2022; Pisner et al., 2019; Satyshur et al., 2018) and rumination-related dynamic FC (Chen (••) & Yan (•超•), 2021; Kaiser et al., 2016; Kucyi & Davis, 2014) alterations; and(2) increase in brooding severity would be significantly associated with decrease in time spent and increased temporal variability in the dFNC states during brooding rather than resting-state, as experimentally-induced brooding is expected to be more sensitive in capturing the active cognitive aspect of brooding compared to resting-state (Berman et al., 2014; Chen (••) & Yan (•超•), 2021; Misaki et al., 2023). Since our clinical trial did not include a brooding condition after neurofeedback, we performed an exploratory analysis to identify any changes in the dynamics of brooding-related dFNC states from pre-to post-neurofeedback resting-state.

## Methods

The study protocol was approved by the Western Institutional Review Board (IRB#20210286) and registered on ClinicalTrials.gov (NCT04941066). The complete study details can be found elsewhere (Tsuchiyagaito et al., 2021, 2023).

### Study sample

The recruited subjects comprised 39 individuals with MDD and 28 healthy control (HC) volunteers. All subjects were aged 18-65 years, fluent in English, and did not endorse any abnormal neuromorphological brain profiles, pregnancies or contraindications to MRI. MDD inclusion criteria involved: unipolar MDD categorized by Mini-International Neuropsychiatric Interview 7.0.2 (MINI) (Sheehan et al., 1998) and Montgomery-Åsberg Depression Rating Scale (MADRS) scores > 6 (Montgomery & Asberg, 1979). MDD exclusion criteria included: lifetime history of bipolar disorder, schizophrenia or other psychotic disorders, DSM-5 criteria for substance abuse or dependence within 6 months before study screening, active suicidal ideation measured using Columbia-Suicide Severity Rating Scale (C-SSRS) (Posner et al., 2011), suicide attempts within 12 months before study screening, commencement of psychotropic medication for depression and/or anxiety less than a month before study screening, or commencement of psychological therapy less than a month before study screening. HC volunteers had no prior psychotropic medication use or neuropsychiatric conditions as assessed by MINI. All participants provided written informed consent and received financial compensation for their participation.

### MRI scanning

MRI data was acquired on a 3 Tesla MR750 Discovery (GE Healthcare) scanner with 8-channel receive-only head array coil. Blood-oxygen-level-dependent (BOLD) fMRI data was acquired using T2*-weighted gradient echo-planar sequence with sensitivity encoding (GE-EPI SENSE) which had the following parameters: TR/TE=2000/25ms, acquisition matrix=96×96, FOV/slice=240/2.9mm, flip angle=90°, voxel size 2.5×2.5×2.9mm^3^; 40 axial slices, SENSE acceleration R=2. The anatomical T1-weighted (T1w) MRI images were acquired using magnetization-prepared rapid gradient-echo (MPRAGE) sequence with parameters: FOV=240×192 mm, matrix=256×256, 124 axial slices, slice thickness=1.2 mm, 0.94×0.94×1.2mm^3^ voxel volume, TR/TE=5/2ms, SENSE acceleration R=2, flip angle=8°. Concurrent physiological signals were recorded using MRI-compatible GE respiration belt and pulse oximetry sensor.

### FMRI design

The fMRI session included resting-state (6min 50s), experimentally induced brooding condition (6min 50s), 3 neurofeedback runs with baseline and transfer, and post-neurofeedback resting-state (6min 50s). During resting-state, subjects were instructed to clear their mind and not think about anything in particular. Prior to MRI, all subjects were instructed to specify emotionally salient personal events that triggered brooding (e.g., experiencing rejection). These events were subsequently used as personalized cues to elicit brooding in the scanner, with a specific instruction, *‘Why did you react the way you do?”* while subjects viewed a fixation cross. For neurofeedback, subjects were instructed to implement strategies for regulating brooding, guided by real-time visual feedback associated with decrease in their FC between PCC and right TPJ. Each of the MDD and HC groups were further subdivided into an active group (MDD N=18; HC N=13) receiving contingent and real FC neurofeedback, and a sham group (MDD N=18; HC N=13) receiving non-contingent and artificially synthesized neurofeedback.

### Brooding score

Here, we examined the relationship between the intensity of brooding, measured using the ‘brooding’ subscale of the 22-item Ruminative Response Scale (RRS-B) (Nolen-Hoeksema & Morrow, 1991; Treynor et al., 2003), and dynamic FNC. With a 4-point Likert scale, RRS-B evaluates one’s tendency to passively dwell on causes and consequences of depressive events/mood (e.g., *think* ‘*why can’t I handle things better’*).

### MRI preprocessing

Data was converted to BIDS format and preprocessed using fMRIPrep 23.1.3. T1-weighted (T1w) anatomical images were corrected for intensity non-uniformity, and skull-stripped using Advanced Normalization Tools (ANTs) (Avants et al., 2009). Brain tissue segmentation of cerebrospinal fluid (CSF), white matter (WM) and gray matter (GM) was performed on the brain-extracted T1w using FSL FAST (Smith et al., 2004; Zhang et al., 2001). Finally, volume-based spatial normalization of the brain-extracted T1w images to standard space (MNI152NLin2009cAsym) was performed through nonlinear registration using ANTs.

Prior to preprocessing fMRI BOLD data, its first 5 volumes were discarded to allow for equilibration of the magnetic fields, resulting in 200 volumes per run per subject. Subsequently for each run and subject, the following preprocessing steps were performed in fMRIPrep (Esteban et al., 2019). Reference volume and skull-stripped versions of the BOLD data were generated. The BOLD reference was co-registered to the T1w reference using boundary-based registration with six degrees of freedom in FreeSurfer (Greve & Fischl, 2009). Head-motion parameters with respect to the BOLD reference (transformation matrices, and six rotation and translation parameters) were estimated prior to spatiotemporal filtering. Fieldmap-less B0 inhomogeneity distortion correction was performed by fMRIPrep, and slice-timing correction was performed using AFNI (Cox & Hyde, 1997). All resamplings were performed with a single interpolation step to derive the fully preprocessed spatially normalized BOLD data. Spatial smoothing was performed on the pre-processed data using a gaussian kernel size of 6 mm full-width half-maximum with FSL. Denoising with nuisance regression was performed separately on the outputs of ICA prior to estimation of dFNC. Physiological nuisance predictors (8 respiration, 6 cardiac, 4 respiration x cardiac, 1 heart rate (Chang et al., 2009), 1 respiratory volume (Harrison et al., 2021)) were estimated with RETROspective Image CORrection (RETROICOR) (Glover et al., 2000) within the PhysIO toolbox (Kasper et al., 2017) using the respiration and cardiac data measured during fMRI.

Subjects whose mean framewise displacement (mean FD; estimated in fMRIPrep) exceeded a threshold of 0.3 mm were excluded (3 MDD subjects and 2 HCs) due to excessive head motion, thereby leading to a final sample of 36 MDD subjects and 26 HCs.

### Independent Component Analysis (ICA)

Following standard recommendations (Allen et al., 2014) in GIFT toolbox v4.0.4.10 (https://trendscenter.org/software/gift/), the preprocessed BOLD data was combined across all subjects (from MDD and HC groups) and runs, decomposed into functional networks using group-level spatial ICA, and denoised. Specifically, after intensity normalization of the preprocessed data, dimensionality reduction was initiated via subject-level principal components analysis (PCA) (130 PCs). Subsequently, group-level PCA (across all runs and subjects) using expectation-maximization (EM) algorithm (Roweis, 1997) retained 100 PCs. The Infomax ICA algorithm (Bell & Sejnowski, 1995) was repeated 15 times in ICASSO (http://www.cis.hut.fi/projects/ica/icasso/) to estimate 100 reliable group ICs. Subject-specific ICs were derived from the group ICs using GICA1 back-reconstruction (Calhoun et al., 2001; Erhardt et al., 2011). Following established guidelines for IC classification (Griffanti et al., 2017), two raters (SG and AT) independently classified the 100 group ICs into signal and artifactual (e.g., physiological, movement, imaging artifacts) ICs based on spatial, temporal and spectral characteristics. Final consensus between raters enabled identification of 34 signal ICs (Figure 2) as functional networks showing peak activations in known cortical and sub-cortical regions, minimal spatial overlap with known vascular, ventricular, motion, and susceptibility artifacts, and predominantly low-frequency time-courses (Allen et al., 2011; Cordes et al., 2000).

All subject-level signal ICs were temporally denoised by low-pass filtering (0.15 Hz cutoff), motion outlier de-spiking (replacing outliers via third-order spline interpolation) (Allen et al., 2014), detrending linear, quadratic and cubic trends, and multiple regression using 20 RETROICOR physiological and 12 fMRIPrep head motion (6 rotation+translation & derivatives) regressors.

### DFNC estimation

DFNC was estimated with standard settings in the temporal dFNC toolbox (Allen et al., 2014) packaged within GIFT. Specifically, sliding window covariance (window length=22TRs(44s), Gaussian taper σ=3TRs, step length=1TR) was computed across the 34 denoised IC timecourses, resulting in 178 concatenated sequential FNC windows per run per subject. Covariance was computed from sparse L1-regularized precision matrices (Smith et al., 2011; Varoquaux et al., 2010) using a graphical LASSO approach (Friedman et al., 2008), wherein the regularization parameter lambda (λ) was optimized via within-run cross-validation framework. The dFNC estimates were controlled for subject-level covariates including age, sex and mean FD, and Fisher transformed resulting in normalized correlation matrices (34×34) that varied across time for each run per subject.

To investigate the dynamics of recurring FNC states, k-means clustering (Lloyd, 1982) (with Manhattan distance) was performed on the windowed correlation matrices. Initial clustering with randomized centroid initializations was performed 500 times on subsampled data (subject exemplars (Pascual-Marqui et al., 1995)) to avoid local minima while minimizing computational burden. The resulting centroids (cluster medians) were then used to initialize clustering of all data (62subjects x 3runs x 178windows=33,108matrices). The optimal number of clusters was determined as four (k=4) using the elbow criterion of the cluster validity index (within-cluster distance ⎟ between-cluster distance).

### Outcomes

Based on the dFNC state transitions of each fMRI condition and subject (Figure 1), dwell time and fraction of time were calculated for each of the four dFNC states. Dwell time refers to the average number of consecutive FNC windows occupied by a given dFNC state. Fraction of time represents the proportion of total time spent in a given dFNC state. The dwell time and fraction of time of each dFNC state were compared between MDD and HC during the baseline resting-state and the brooding condition using independent non-parametric Wilcoxon rank sum tests. Subsequently, the association between these outcomes (dwell time, fraction of time) and brooding (RRS-B scores) were examined using non-parametric Spearman correlation for each dFNC state during the baseline resting-state and brooding condition, within the MDD and HC groups separately. FDR correction (p<0.05) was used to control for the multiple comparisons. Note that not all subjects visit every dFNC state during an fMRI run. Therefore, we also conducted a chi-squared test of proportions to examine if there were any significant differences between MDD and HC in the proportion of subjects entering each dFNC state, during resting-state and brooding condition.

**Figure 1:**
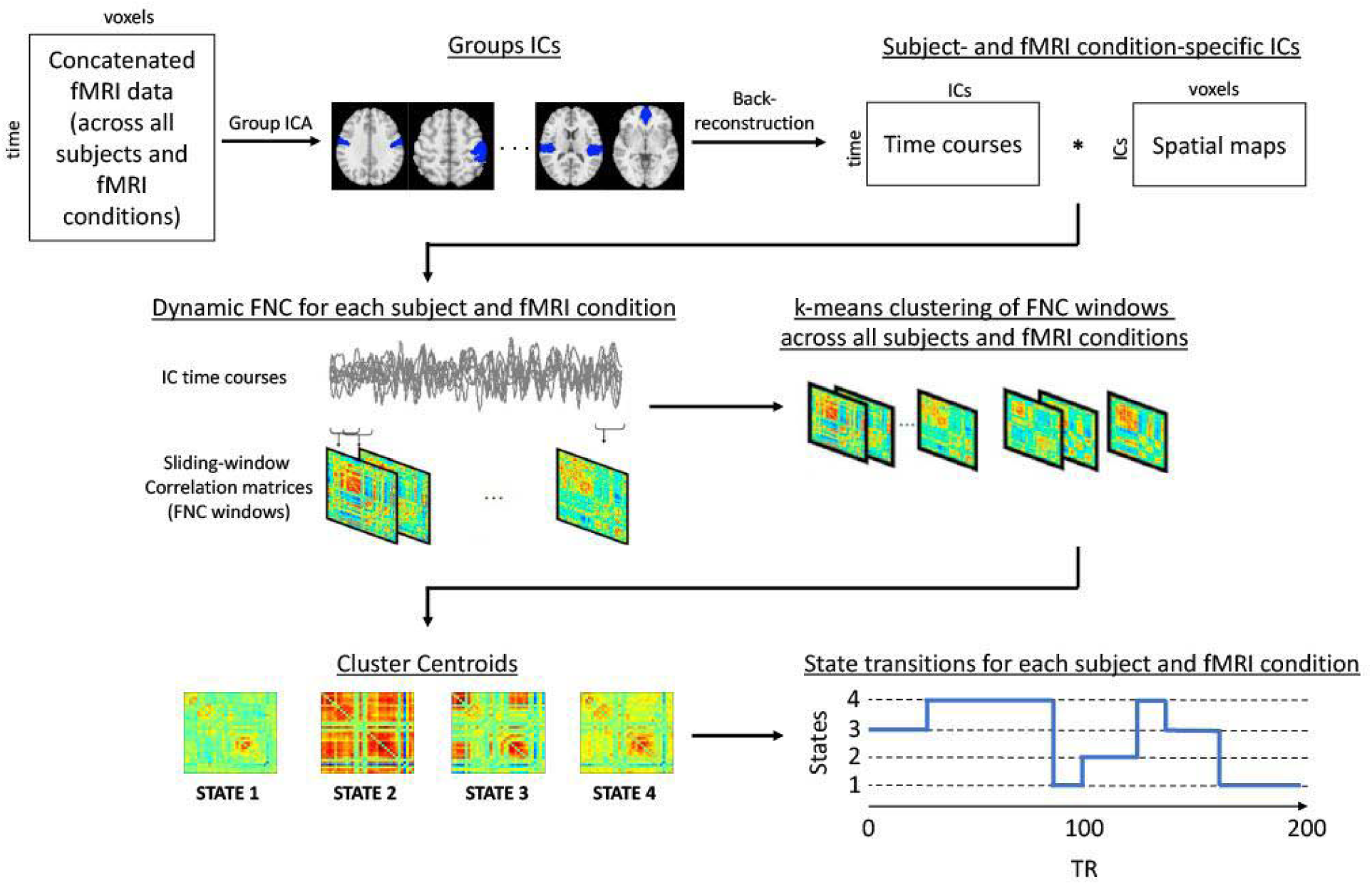
Graphical illustration of the dynamic Functional Network Connectivity (dFNC) method used in the study. Group Independent Component Analysis (ICA) is performed on concatenated data and subsequent back-reconstruction produces subject-specific and fMRI condition-specific independent components (ICs). Sliding-window correlation is performed across the timecourses of these ICs, to extract FNC matrices that are then clustered to produce group-level centroid states. Finally, the state transitions are estimated for each subject and fMRI condition, which are used to compute mean dwell time and fraction of time associated with each centroid state.

As a secondary analysis, non-parametric Wilcoxon signed rank tests were conducted to explore the difference between time spent (i.e., dwell and fraction) in RRS-B related dFNC state/(s) during baseline resting-state and post-neurofeedback resting-state within each neurofeedback subgroup. This was to examine if and whether time spent in the dFNC state/(s) related to brooding was affected by the neurofeedback training.

## Results

The final sample included HC (N=26) and MDD (N=36) groups each subdivided into active and sham neurofeedback subgroups. Table 1 lists the sample size, age, sex and RRS-B scores in each subgroup.

**Table 1:** Age, sex, sample size and baseline RRS-B scores in MDD active, MDD sham, HC active and HC sham groups.

| Sample type | Sample size (N) | Age in years (mean±s.d) | Females/Males | RRS-B scores (mean±s.d) |
| --- | --- | --- | --- | --- |
| MDD active | 18 | 32.2±10.1 | 12/6 | 12.9±3.2 |
| MDD sham | 18 | 32.4±10.9 | 14/4 | 12.5±3.2 |
| HC active | 13 | 23.2±4.3 | 9/4 | 6±1.2 |
| HC sham | 13 | 22.3±3.7 | 11/2 | 6.4±1.1 |

### Functional networks from ICA

A total of 34 functional networks (ICs) were identified from the group ICA analysis. These networks were labeled and grouped into six domains based on standard taxonomy (Uddin et al., 2019), and established network (Yeo et al., 2011) and subcortical (Tian et al., 2020) parcellations. The domains include default-mode, central executive, salience, attention, somatomotor, and visual. Two cortical networks and the five subcortical networks did not fit into a single domain. Figure 2 shows the spatial brain maps and labels of all the functional networks identified using ICA, grouped within their respective domains.

**Figure 2:**
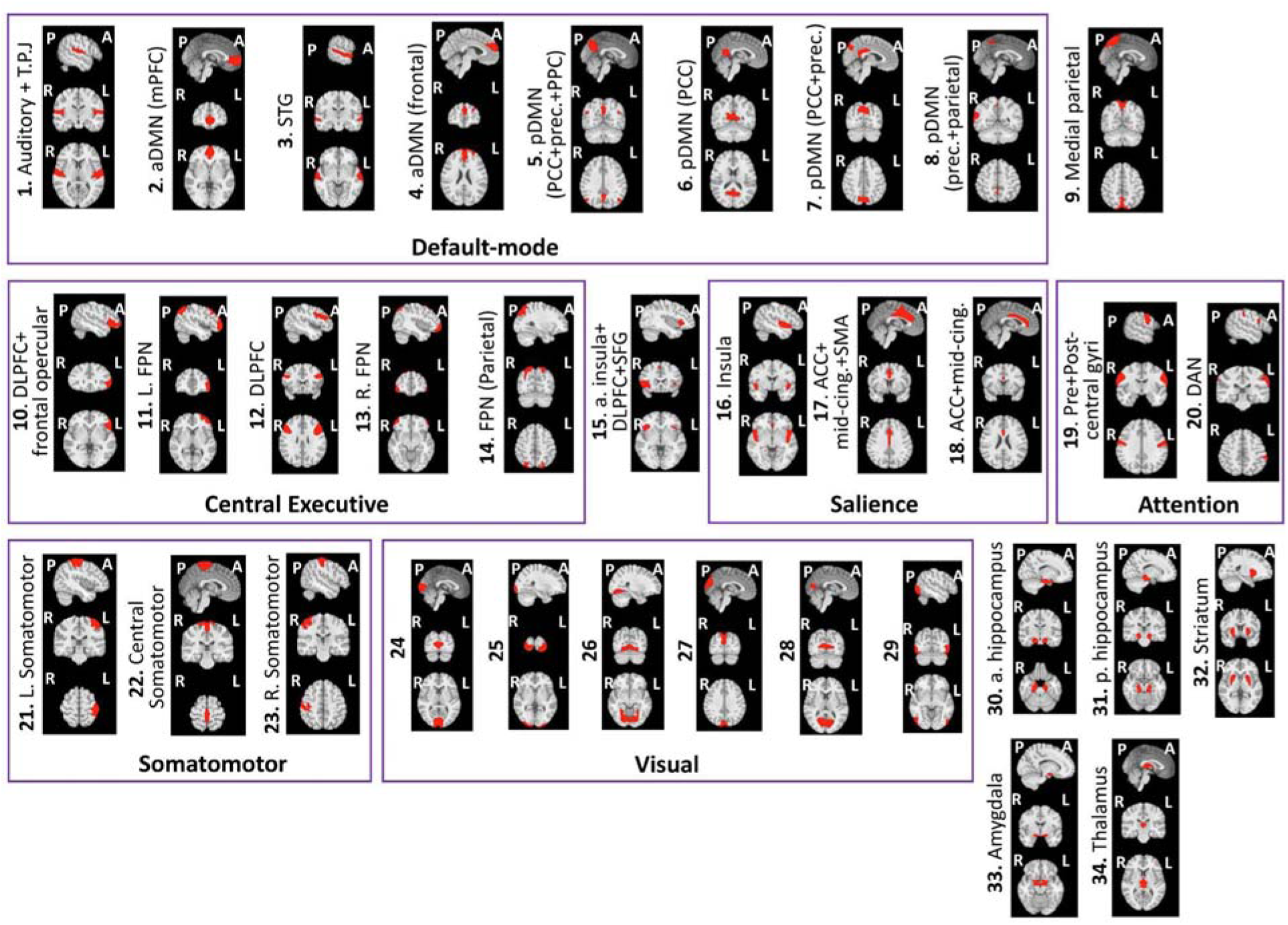
Spatial maps of the 34 functional networks extracted from group ICA, overlaid on discrete anatomical brain slices. Where applicable, the networks are labeled and grouped within rectangles into their respective domains of functional affiliation. Note that functional networks 24-29 indicate different visual subnetworks. A – anterior, P – posterior, R – Right, L – Left; T.P.J - Temporoparietal junction, aDMN - anterior default-mode network, pDMN - posterior default-mode network, mPFC - medial prefrontal cortex, STG - superior temporal gyrus, prec. - precuneus, PCC - posterior cingulate cortex, PPC - posterior parietal cortex, DLPFC - dorsolateral prefrontal cortex, FPN - frontoparietal network, SFG - superior frontal gyrus, ACC - anterior cingulate cortex, mid-cing. - mid-cingulate cortex, SMA - supplementary motor area, DAN - dorsal attention network, a. hippocampus - anterior hippocampus, p. hippocampus - posterior hippocampus.

### Dynamic functional network connectivity (dFNC) states

Clustering of the time-varying FNC states formed by the 34 functional networks produced four centroid dFNC states, as shown in Figure 3.

**Figure 3:**
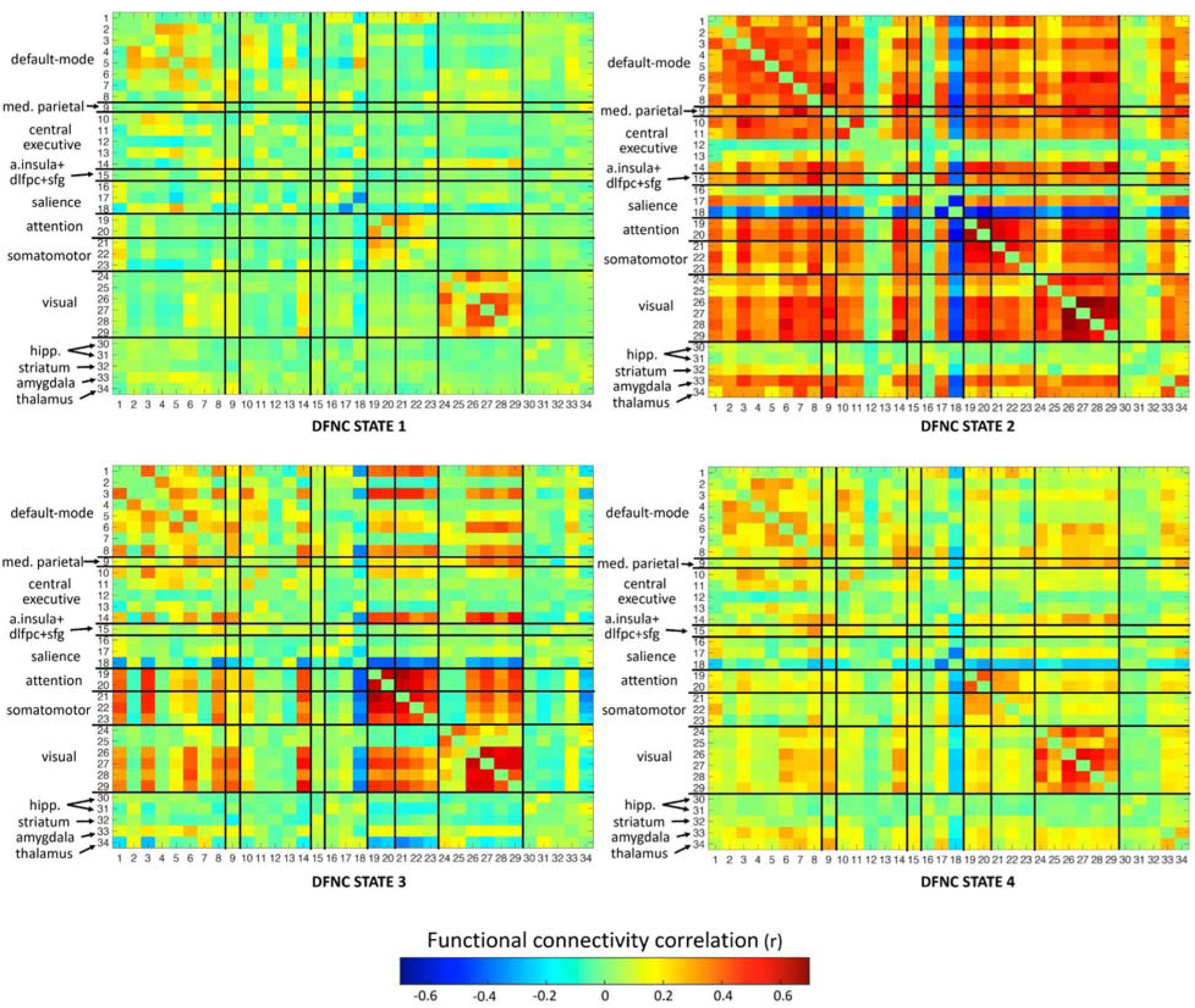
Graphical representation of the four recurring centroid dFNC states extracted from dFNC analysis. The matrices are symmetric, and the black lines within each matrix indicate boundaries of functional network domains (as displayed in Figure 2). The value of functional connectivity correlation between any two IC networks determines the color (‘hot’ color gradient) of the corresponding matrix entry. med. - medial, a. insula - anterior insula, dlpfc - dorsolateral prefrontal cortex, sfg - superior frontal gyrus, hipp. - hippocampus.

DFNC state 1 is a sparsely connected state, marked by moderate positive FC within default-mode (between anterior and posterior default-mode networks), and dorsal attention networks, strong positive FC within visual networks, and strong negative FC within anterior and mid-cingulate salience networks. DFNC state 2 is a hyperconnected state, with strong positive FC within and between most networks throughout the brain, and strong negative FC between the anterior and mid-cingulate salience network and the whole brain. The hippocampi, striatum and some central executive subnetworks however show weak FC with the whole brain.

DFNC state 3 is a densely connected integrated state, comprising strong positive FC within attention, somatomotor and visual networks, moderate-to-strong positive FC within default-mode networks, and strong positive FC between attention, somatomotor, default-mode (superior temporal gyrus (STG) and temporoparietal junction (TPJ)) and visual networks. Posterior default-mode networks (posterior cingulate cortex (PCC) and precuneus) show strong positive FC with visual networks. The thalamus shows strong negative FC with attention, somatomotor and default-mode networks (TPJ and STG). The anterior and mid-cingulate salience network shows strong negative FC with somatomotor, attention, visual, central executive parietal, medial parietal, and default-mode networks (TPJ, STG and precuneus). Subnetworks from CEN (DLPFC, parietal CEN) have moderate-to-strong positive FC with attention, visual, somatomotor and default-mode networks.

DFNC state 4 is characterized by strong positive FC within visual and attention networks, moderate-to-strong positive FC within default-mode networks, scattered moderate positive FC of default-mode with visual, central executive with default-mode, attention with somatomotor, amygdala and thalamus with default-mode, and parietal with visual and default-mode networks, and moderate-to-strong negative FC between anterior/mid-cingulate network and the whole brain.

### Differences in time spent in dFNC states between groups

There were no significant differences in the time spent (dwell time or fraction of time) in any dFNC state between MDD and HC groups during baseline brooding condition or resting-state. Mean values of dwell time and fraction of time for each condition, group and dFNC state can be found in Supplementary Table 1. Additionally, the proportion of subjects entering each dFNC state was not significantly different between MDD and HC groups during resting-state or brooding. The proportions of subjects per dFNC state for each group and condition are shown in Supplementary Table 2.

### Association between time spent in dFNC states and RRS-B scores

The dwell time and fraction of time spent in dFNC state 3 showed strong negative correlation with RRS-B scores in the MDD group during the brooding condition. These were the only associations that remained significant after FDR correction across all correlation analyses (dwell time: r(34)=-0.5, p-FDR=0.031; fraction of time: r(34)=-0.5, p-FDR=0.031) (Figure 4a, 4b). The correlations were non-significant for MDD in resting-state (Figure 4e, 4f) and for the HC group (Figure 4c, 4d, 4g, 4h). Illustrations of the correlations for the other dFNC states are shown in Supplementary Figures SF1, SF2 and SF3. None of these survived FDR correction.

**Figure 4:**
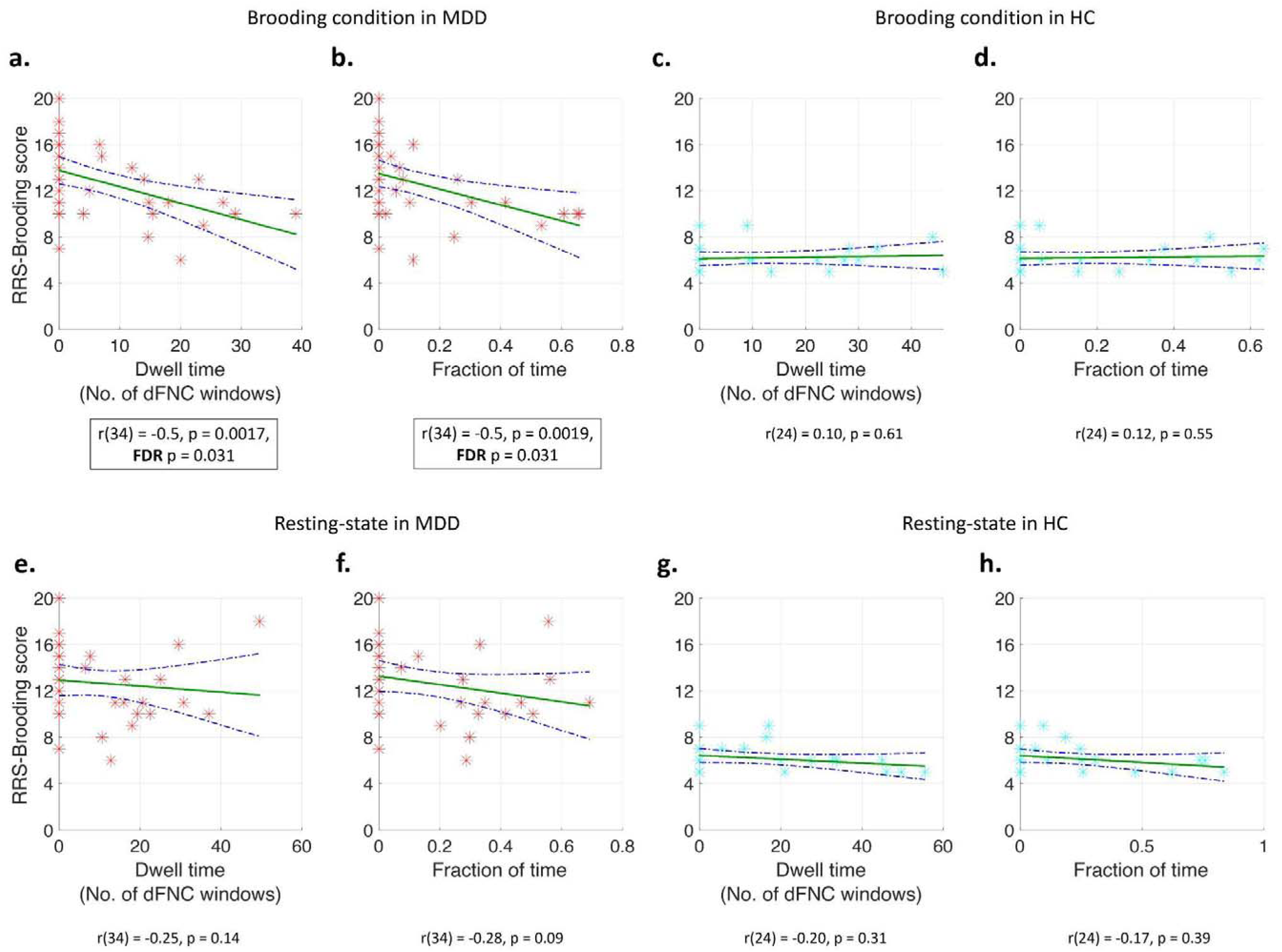
Scatter plots showing associations between brooding severity (RRS-B scores) and outcomes of **dFNC state 3** in MDD group during brooding condition (dwell time in (a) and fraction of time in (b)), HC group during brooding condition (dwell time in (c) and fraction of time in (d)), MDD group during resting-state (dwell time in (e) and fraction of time in (f)), and HC group during resting-state (dwell time in (c) and fraction of time in (d)). In each scatter plot, the RRS-B scores are depicted in the y-axis while the outcome of dFNC state 3 is shown in the x-axis. The green line represents the linear fit of the association between RRS-B scores and the dFNC outcome, while the blue dotted curved lines represent the 95% confidence interval of the linear fit. The only associations significant after FDR correction for multiple comparisons were observed in the MDD group during the brooding condition ((a) and (b)). RRS-B - Rumination Response Scale - Brooding subscale

### Differences in time spent in dFNC state 3 between pre-neurofeedback and post-neurofeedback resting-state

Neurofeedback training related changes in dwell time and fraction of time spent in the brooding-associated dFNC state 3 in MDD active, MDD sham, HC active and HC sham subgroups were explored through paired t-tests (Figure 5). The fraction of time spent in dFNC state 3 showed a significant increase from pre-neurofeedback resting-state to post-neurofeedback resting state in the MDD active neurofeedback group only (Figure 5e; z=-2.09, p=0.037). Changes in dFNC state 3 measures were non-significant (Figure 5) for all other neurofeedback subgroups (i.e., HC active, HC sham and MDD sham).

**Figure 5:**
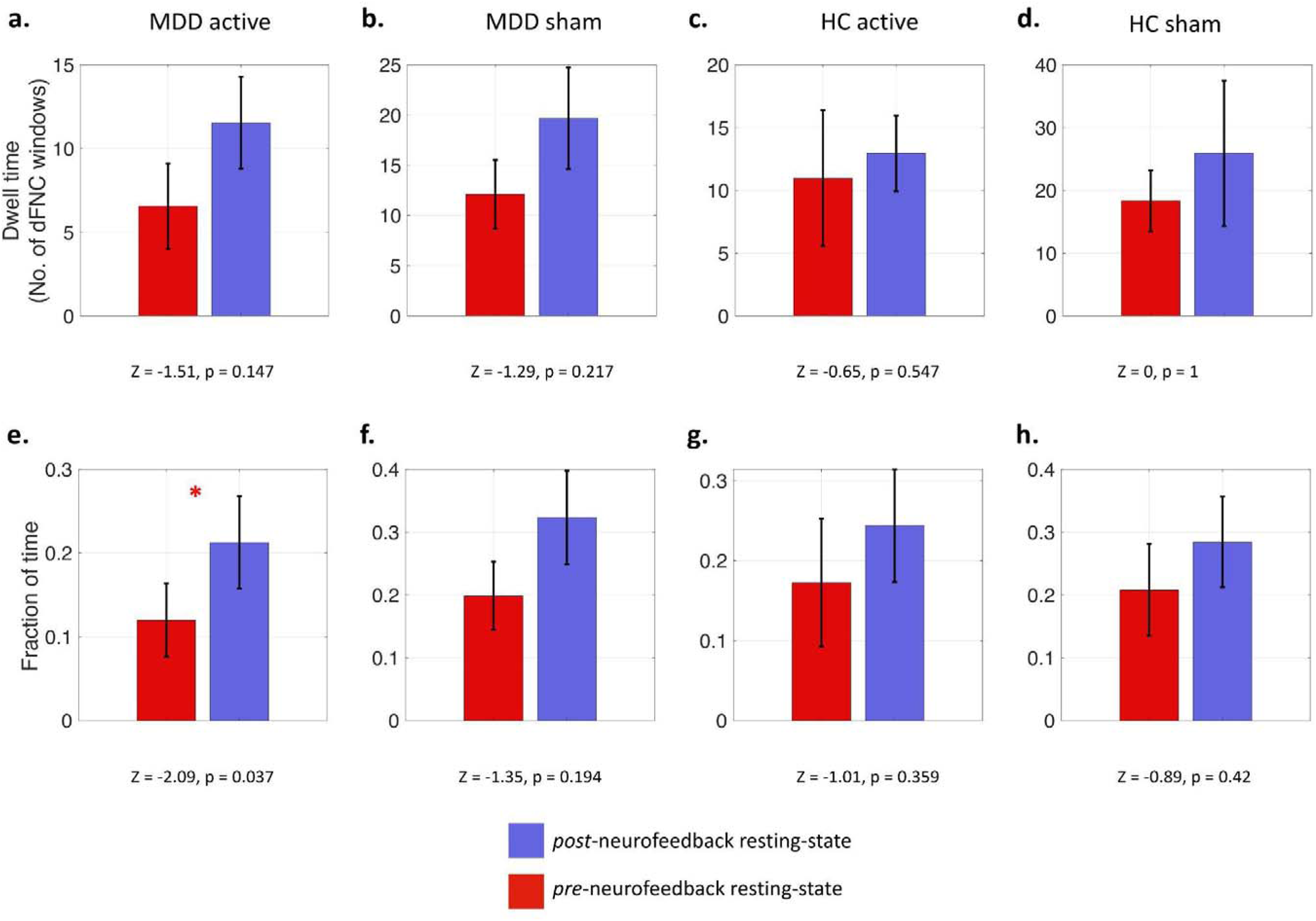
Bar graphs, with corresponding p values and z-score approximations, representing mean changes in the outcomes of **dFNC state 3** from pre-to post-neurofeedback resting-state in the MDD active neurofeedback ((a) and (e)), MDD sham neurofeedback ((b) and (f)), HC active neurofeedback ((c) and (g)), and HC sham neurofeedback ((d) and (h)) subgroups. The top four graphs ((a)-(d)) represent changes in dwell time of dFNC state 3 (y-axis), while the bottom four graphs ((e)-(h)) represent changes in fraction of time of dFNC state 3 (y-axis). The bars represent mean values and the black lines on each bar represents standard error about the mean. The only significant change (indicated by red asterisk) was found in the MDD active neurofeedback group (e), where the fraction of time spent in dFNC state 3 increased after neurofeedback. This significance however did not survive correction for multiple comparisons.

## Discussion

We examined whole-brain time-varying dynamic functional network connectivity (dFNC) associated with resting-state and brooding fMRI in depressed and healthy individuals to illuminate brain states associated with brooding severity, a critical symptom of depression measured using RRS-B scores. We identified four group-level summary dFNC states that were inhabited for varying durations by each individual in each fMRI condition. The first hypothesis, positing that the time spent in the identified dFNC states would differ between MDD and HC during resting-state and brooding conditions, was not supported. Time spent in these states was not significantly different between MDD and HC in resting-state or the brooding condition, suggesting that the presence and maintenance of these states are not uniquely altered in MDD at a detectable level with the current sample size. However, our second hypothesis was supported: greater brooding severity was significantly associated (FDR-corrected p<0.05) with lesser time spent (i.e., proportion of time and dwell time) in the densely connected dFNC state 3, which is primarily characterized by moderate-to-strong positive FC within and between default-mode, attention, somatomotor, and visual networks, between central-executive and default-mode regions, and strong negative FC of ACC and thalamus with aforementioned networks. Notably, this relationship was significant only in the MDD group during the brooding condition, highlighting the utility of mood induction paradigms in capturing neurobiological effects sensitive to brooding and rumination in MDD.

Secondary analysis further revealed that our real-time fMRI neurofeedback trial was associated with significant increase (uncorrected p=0.037) in the proportion of time spent in the anti-brooding dFNC state 3. The increase from pre-to-post neurofeedback resting-state was significant only in the MDD active neurofeedback group. Such preliminary evidence suggests that neurofeedback may mitigate brooding in MDD beyond sham neurofeedback by modifying brooding-related FC dynamics, in addition to time-averaged FC as previously demonstrated (Tsuchiyagaito et al., 2023).

### Dynamic FC associated with brooding in depression

Consistent with our hypothesis, brooding was associated with time spent in a densely connected dFNC state 3 containing unique FC patterns involving DMN, SN (ACC), CEN (parietal areas) and subcortical areas (thalamus). We also found prominent involvement by additional networks, namely dorsal attention, somatomotor and visual. DFNC state 3 is an integrated densely connected state comprising moderate-to-strong FC primarily within and between default-mode, attention, somatomotor and visual networks. The temporal dynamics of these connections are particularly relevant to MDD. MDD has been associated with more time spent having reduced FC within somatomotor and dorsal attention networks (Javaheripour et al., 2023), less time in integrated states with increased FC between sensory and default-mode networks (Wu et al., 2019), and more time with reduced FC within and between visual, auditory, somatomotor and default-mode networks (Xu et al., 2022). Similarly, recent mega- and meta-analytic evidence of prominent static FC alterations in MDD implicated hypoconnectivity within and between dorsal attention, somatomotor, parietal and visual networks (Javaheripour et al., 2021; Tse et al., 2023). This is consistent with our observation of increased MDD brooding severity with decrease in time spent having hyperconnectivity in these same networks. Importantly, as shown in Figure 4, several MDD individuals with high brooding severity did not even visit the densely connected dFNC state 3 (i.e., fraction of time = 0).

Higher interconnectedness of the default-mode network with the attention, somatomotor and visual networks, as observed in dFNC state 3, may facilitate improved integration of ongoing self-referential processing with present-moment environmental, sensory and bodily experiences. Such improved integration can potentially enable frequent interruptions to passive brooding in MDD through attention to present-moment stimuli triggering mood change. DFNC state 3 additionally includes strong negative FC involving mid-cingulate cortex, ACC and thalamus. Mid- and anterior cingulate cortices are generally implicated in emotion and cognitive regulation (Stevens et al., 2011), while thalamus is involved in brain-wide, multimodal sensory processing, arousal and perception (Hwang et al., 2017; Shine et al., 2023). These areas are particularly dysfunctional in MDD, brooding and rumination (Berman et al., 2011; Chen (••) & Yan (•超•), 2021; Long et al., 2020; Yao et al., 2019). Therefore, their strong anticorrelation with auditory default-mode (TPJ, STG), somatomotor, attention and visual networks in dFNC state 3 suggests that more time in this state may promote adaptive emotional and cognitive processing, facilitated by increased integration of somatosensory and external perceptual updates into self-related thinking. Additionally, such adaptive processing is likely facilitated by the state’s moderate-to-strong positive FC between CEN and DMN regions, which may promote cognitive disengagement from brooding in MDD (Y. Li et al., 2021; Pisner et al., 2019).

On the contrary, shorter fraction of time and dwell time (i.e., time spent continuously in a state before transitioning to another state) in dFNC state 3 or not visiting the state at all likely minimize these adaptive processes, exacerbating brooding and MDD. This effect is consistent with several dynamic FC studies that found increases in rumination with higher temporal variability of FC in several areas and networks, including DMN, visual network, somatosensory network, temporal areas, mPFC, ACC, dorsal attention network, inferior parietal lobe (IPL), TPJ and superior parietal lobe (SPL) (Chen (••) & Yan (•超•), 2021; Kim et al., 2023; Kucyi & Davis, 2014). Particularly, these areas also form prominent FC patterns in dFNC state 3, whose shorter dwell time (or higher temporal variability) is related to increased brooding severity here, illustrating some of the shared neural dynamics underpinning rumination and its MDD-sensitive component - brooding. Shorter dwell times in strongly connected states similar to dFNC state 3 have also been associated with suicide risk and ideation (Xu et al., 2022).

Dynamic time-varying FC is a promising avenue to elucidate intricate time-sensitive neuronal mechanisms underlying various states of cognition, disease and consciousness, with greater potential to characterize psychiatric biomarkers compared to traditional static FC alone (Calhoun et al., 2008, 2014; Cohen, 2018; Ganesan et al., 2022; Hutchison et al., 2013). Notably, dynamic FC can outperform static FC in predicting individual differences in rumination using diverse clinical and subclinical datasets (Kim et al., 2023). Our present work demonstrates the utility of dynamic FC in characterizing the time-varying behavior of a whole-brain FC state associated with brooding, a critical symptom and prognostic factor of MDD. In addition to unique FC patterns comprising DMN, CEN, SN and subcortical networks, we found prominent involvement by somatomotor, attention and visual networks in brooding-related FC dynamics.

### Effect of real-time neurofeedback on brooding-related dynamic FC in depression

Our secondary analysis also highlighted the sensitivity of dynamic FC in capturing intervention effects associated with real-time fMRI neurofeedback. Specifically, following real-time neurofeedback training aimed at attenuating brooding, MDD subjects were able to spend significantly more time in dFNC state 3 during rest, suggesting diminished brooding. Importantly, this effect was non-significant in the sham neurofeedback MDD group that received artificially synthesized feedback signals, and in the HC subgroups. This suggests a neurobiologically adaptive response to the neurofeedback training in MDD, indicating reduced difficulty in sustaining the protective dFNC state 3 and facilitating a move away from passive dwelling on maladaptive thought patterns. This is also consistent with previous findings (Tsuchiyagaito et al., 2023) which showed that only the MDD active neurofeedback, and not MDD sham neurofeedback, subgroup experienced significant reduction in brooding severity measured one week after neurofeedback. Overall, the current work highlights the utility of time-varying FC in examining neurofeedback-related outcomes to capture effects that may be missed by traditional static FC approaches.

### Limitations

The sample size used in this study was small and imbalanced between MDD (N=36) and HC (N=26) groups. This may have biased the dFNC clustering process towards MDD-related states. Although a modest proportion of subjects did not visit dFNC states 2 and 3 during rest or brooding (i.e., fraction of time = 0), these proportions were similar in both groups suggesting that such densely connected dFNC states may be occupied less commonly in general. To increase generalizability, our MDD inclusion criteria did not consider the dosage and duration of antidepressant medication use, which could have impacted our findings. Brain areas relevant to MDD such as cerebellum (Phillips et al., 2015), subgenual ACC (Cash et al., 2021), and inferior and polar temporal areas (Berman et al., 2014) were excluded from the whole-brain fMRI analysis, due to issues of MRI signal dropout and limited field-of-view coverage identified during quality assessment. Future studies should examine whole-brain dynamic FC associated with brooding using larger and more balanced samples.

We did not find any significant pre-to-post neurofeedback dFNC changes in the HC groups, likely because of the small sample size and low scope for improvement in brooding from baseline among healthy individuals compared to MDD subjects. Additionally, compared to resting-state, the brooding condition was found to be more sensitive to dFNC changes associated with RRS-B. However, our experimental design did not include a brooding condition post-neurofeedback. Consequently, changes in brooding-related dFNC associated with neurofeedback were examined in resting-state, which may not be as sensitive as mood-induction tasks in capturing neuronal indices of brooding and rumination (Berman et al., 2014; Chen (••) & Yan (•超•), 2021; Misaki et al., 2023), contributing to the weak effects observed in the exploratory neurofeedback analysis. These exploratory findings should hence be interpreted with caution, since the observed significance did not survive correction for multiple comparisons, and group by time interaction effects were not considered due to the small subgroup sizes. Causality cannot be implied, as these findings only suggest potential associations and therapeutic pathways of neurofeedback action on brooding in MDD. Larger fMRI studies with brooding condition pre- and post-neurofeedback, and longitudinal follow-up assessments in the future will help inform the durability of these observed effects on depressive symptomatology and overall cognitive function.

## Conclusion

We investigated whole-brain dynamic functional network connectivity (dFNC) in depressed and healthy individuals during rest and brooding. We found one brain state (dFNC state 3) with distinct FC profiles, whose temporal dynamics were most related to brooding severity, an important symptom and prognostic factor of depression. This dFNC state was densely connected with moderate-to-strong intra- and inter-network FC involving several default-mode, somatomotor, attention, visual, central executive, salience and thalamic areas. Greater time spent and dwell time (stability) in this dFNC state during brooding (but not rest) was significantly associated with lower brooding severity in the major depressive disorder (MDD) group, denoting the state’s potential to offer protection against brooding in MDD. Exploratory analysis revealed that active (and not sham) real-time fMRI neurofeedback targeting PCC-TPJ FC in MDD can potentially increase the time spent in this dFNC state. This work highlights the utility of dynamic FC and mood-induction tasks for investigating fast timescale fluctuations in fMRI brain connectivity patterns associated with a critical symptom of MDD, and presents the promise of real-time fMRI neurofeedback as a tool to cultivate or suppress specific brain states and modulate their dynamics, offering novel insights into personalized non-invasive treatment approaches for depression.

## Data Availability

All data produced in the present study are available upon reasonable request to the authors.

## Acknowledgments

The authors acknowledge Jerzy Bodurka, Ph.D. (1964–2021) for his intellectual and scientific contributions to the establishment of the EEG, structural and functional MRI, and neurofeedback processes at LIBR, that provided the foundation for the data collection, analysis, and interpretation of findings for the present work.

