## Supplementary Material for "Functional brain network dynamics of brooding in depression: insights from real-time fMRI neurofeedback"

*Supplementary Table 1: Measures of time spent (dwell time and fraction of time) per dFNC state by group and baseline fMRI condition*

|  | **Dwell time (no. of dFNC windows) (mean 士 S.D (median))** | | | | **Fraction of time (mean 士 S.D (median))** | | | |
| --- | --- | --- | --- | --- | --- | --- | --- | --- |
|  | **MDD group** | | **HC group** | | **MDD group** | | **HC group** | |
|  | *Baseline Resting-state* | *Brooding condition* | *Baseline Resting-state* | *Brooding condition* | *Baseline Resting-state* | *Brooding condition* | *Baseline Resting-state* | *Brooding condition* |
| **dFNC state 1** | 39.9 士 46.7 (31.0) | 51.5 士 54.7 (29.5) | 52.7 士 61.1 (24.4) | 60.4 士 59.2 (39.7) | 0.51 士 0.33 (0.61) | 0.52 士 0.34 (0.50) | 0.52 士 0.33 (0.52) | 0.55 士 0.34 (0.62) |
| **dFNC state 2** | 11.3 士 30.9 (0) | 10.9 士 28.0 (0) | 6.7 士 9.2  (0) | 6.3 士 11.1 (0) | 0.11 士 0.21 (0) | 0.11 士 0.21 (0) | 0.06 士 0.11 (0) | 0.08 士 0.17 (0) |
| **dFNC state 3** | 9.3 士 12.9 (0) | 7.6 士 10.7 (0) | 14.7 士 18.6 (2.75) | 11.4 士 15.2 (0) | 0.16 士 0.21 (0) | 0.12 士 0.20 (0) | 0.19 士 0.27 (0.03) | 0.16 士 0.23 (0) |
| **dFNC state 4** | 11.7 士 6.7 (12.4) | 13.9 士 11.6 (12) | 12.1 士 8.8 (13.9) | 11.2 士 7.5 (11.8) | 0.21 士 0.15 (0.15) | 0.24 士 0.22 (0.23) | 0.23 士 0.20 (0.18) | 0.20 士 0.17 (0.19) |

*Supplementary Table 2: Proportion of subjects entering each dFNC state by group and baseline fMRI condition.*

|  | **Proportion (in %)** | | | |
| --- | --- | --- | --- | --- |
|  | **MDD group** | | **HC group** | |
|  | *Resting-state* | *Brooding condition* | *Resting-state* | *Brooding condition* |
| **dFNC state 1** | 91.7 | 91.7 | 92.3 | 96.1 |
| **dFNC state 2** | 41.7 | 41.7 | 46.1 | 34.6 |
| **dFNC state 3** | 44.4 | 44.4 | 50.0 | 46.1 |
| **dFNC state 4** | 88.9 | 86.1 | 80.8 | 84.6 |

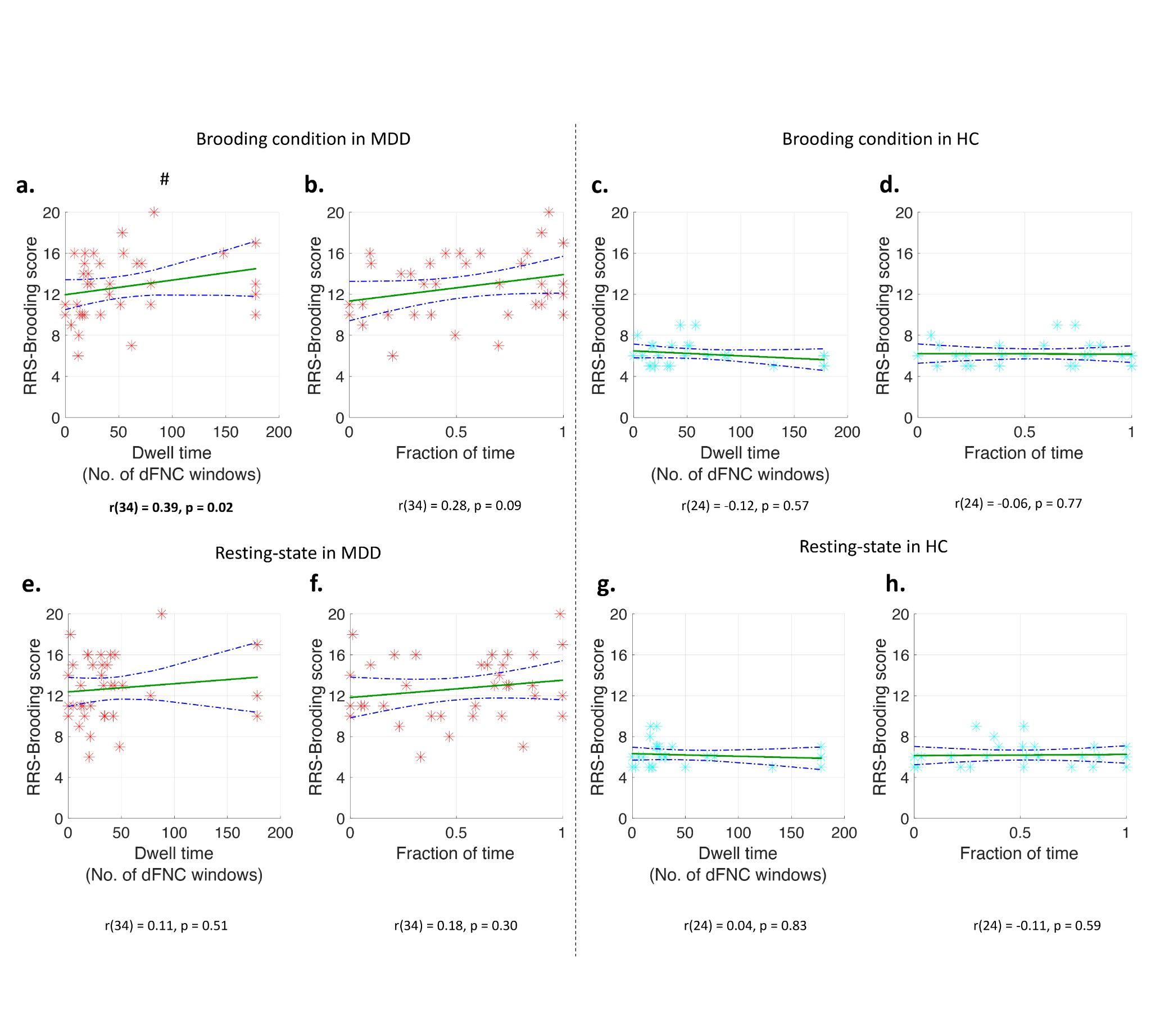

***Figure SF1:*** *Scatter plots showing associations between brooding severity (RRS-B scores) and outcomes of* ***dFNC state 1*** *in MDD group during brooding condition (dwell time in (a) and fraction of time in (b)), HC group during brooding condition (dwell time in (c) and fraction of time in (d)), MDD group during resting-state (dwell time in (e) and fraction of time in (f)), and HC group during resting-state (dwell time in (c) and fraction of time in (d)). In each scatter plot, the RRS-B scores are depicted in the y-axis while the outcome of dFNC state 1 is shown in the x-axis. The green line represents the linear fit of the association between RRS-B scores and the dFNC outcome, while the blue dotted curved lines represent the 95% confidence interval of the linear fit. A hash symbol above a scatter plot indicates that the correlation was significant. Note that none of the correlations were significant after FDR correction for multiple comparisons. RRS-B - Rumination Response Scale - Brooding subscale*

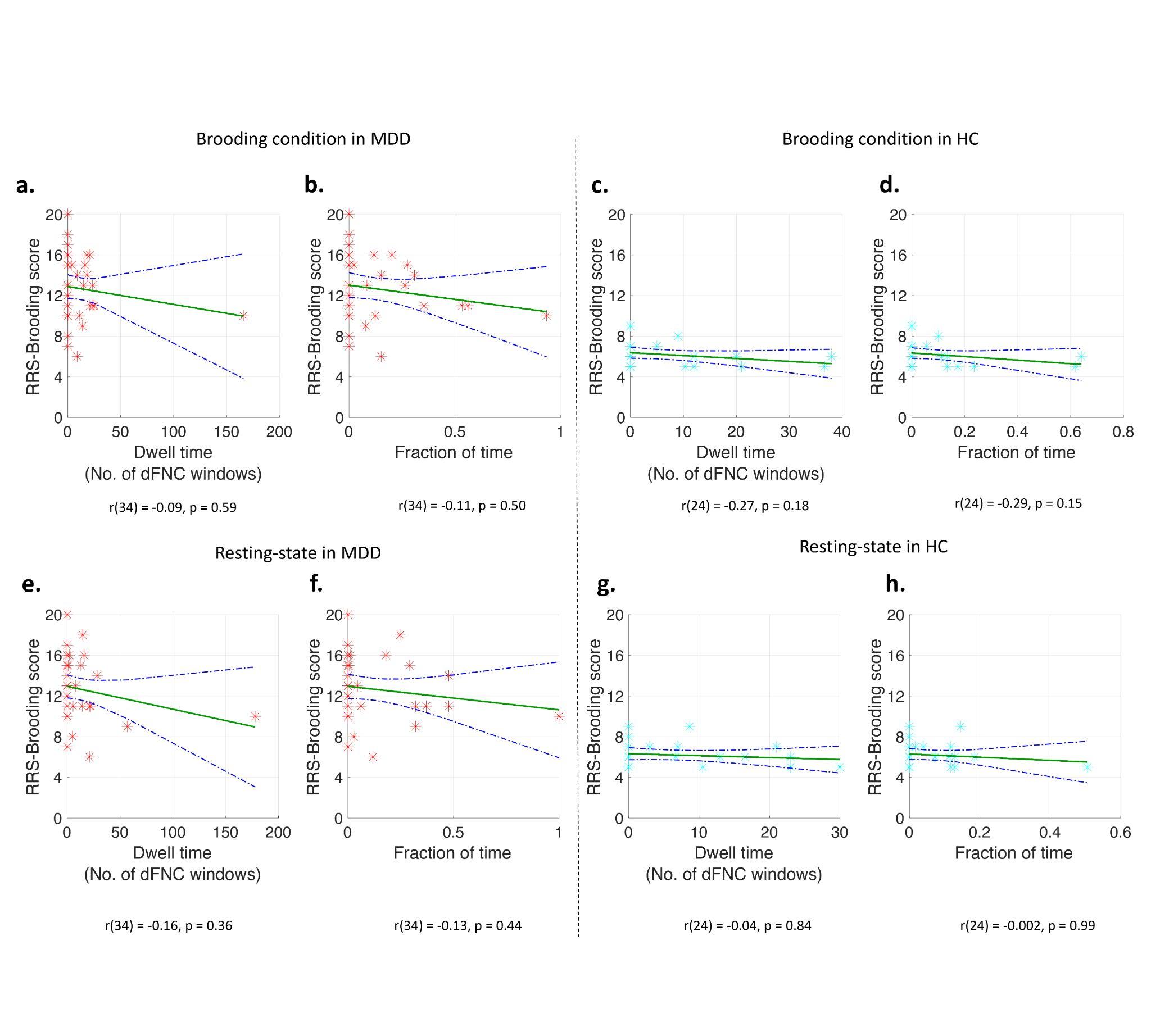

***Figure SF2:*** *Scatter plots showing associations between brooding severity (RRS-B scores) and outcomes of* ***dFNC state 2*** *in MDD group during brooding condition (dwell time in (a) and fraction of time in (b)), HC group during brooding condition (dwell time in (c) and fraction of time in (d)), MDD group during resting-state (dwell time in (e) and fraction of time in (f)), and HC group during resting-state (dwell time in (c) and fraction of time in (d)). In each scatter plot, the RRS-B scores are depicted in the y-axis while the outcome of dFNC state 2 is shown in the x-axis. The green line represents the linear fit of the association between RRS-B scores and the dFNC outcome, while the blue dotted curved lines represent the 95% confidence interval of the linear fit. RRS-B - Rumination Response Scale - Brooding subscale*

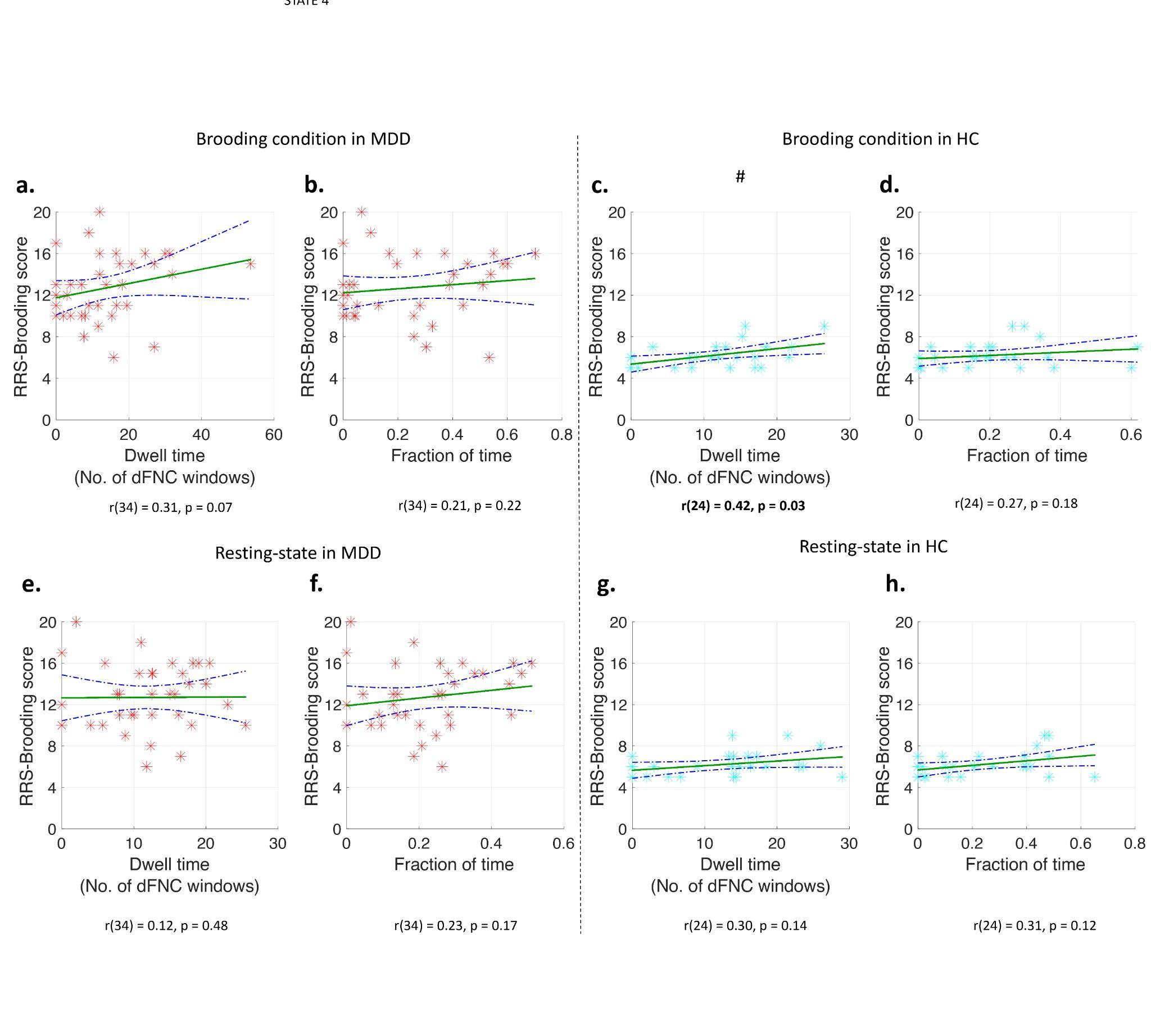

***Figure SF3:*** *Scatter plots showing associations between brooding severity (RRS-B scores) and outcomes of* ***dFNC state 4*** *in MDD group during brooding condition (dwell time in (a) and fraction of time in (b)), HC group during brooding condition (dwell time in (c) and fraction of time in (d)), MDD group during resting-state (dwell time in (e) and fraction of time in (f)), and HC group during resting-state (dwell time in (c) and fraction of time in (d)). In each scatter plot, the RRS-B scores are depicted in the y-axis while the outcome of dFNC state 4 is shown in the x-axis. The green line represents the linear fit of the association between RRS-B scores and the dFNC outcome, while the blue dotted curved lines represent the 95% confidence interval of the linear fit. A hash symbol above a scatter plot indicates that the correlation was significant. Note that none of the correlations were significant after FDR correction for multiple comparisons. RRS-B - Rumination Response Scale - Brooding subscale*
